# Short-Term Corticosteroids in SARS-CoV2 Patients: Hospitalists’ Perspective

**DOI:** 10.1101/2020.06.19.20109173

**Authors:** Vijairam Selvaraj, Kwame Dapaah-Afriyie, Arkadiy Finn, Timothy Flanigan

## Abstract

**Background:** Dexamethasone, a synthetic glucocorticoid, has anti-inflammatory and immunosuppressive properties. There is a hyperinflammatory response involved in the clinical course of patients with pneumonia due to SARS-CoV2. To date, there has been no definite therapy for COVID-19. We reviewed the charts of SARS-CoV2 patients with pneumonia and moderate to severely elevated CRP and worsening hypoxemia who were treated with early, short-term dexamethasone.

**Methods:** We describe a series of 21 patients who tested positive for SARS-CoV2 and were admitted to The Miriam Hospital in Providence and were treated with a short course of dexamethasone, either alone or in addition to current investigative therapies.

**Results:** CRP levels decreased significantly following the start of dexamethasone from mean initial levels of 129.52 to 40.73 mg/L at time of discharge. 71% percent of the patients were discharged home with a mean length of stay of 7.8 days. None of the patients had escalation of care, leading to mechanical ventilation. Two patients were transferred to inpatient hospice facilities on account of persistent hypoxemia, in line with their documented goals of care.

**Conclusions:** A short course of systemic corticosteroids among inpatients with SARS-CoV2 with hypoxic respiratory failure was well tolerated, and most patients had improved outcomes. This limited case series may not offer concrete evidence towards the benefit of corticosteroids in COVID-19. However, patients’ positive response to short-term corticosteroids demonstrates that they may help blunt the severity of inflammation and prevent a severe hyperinflammatory phase, in turn reducing the length of stay, ICU admissions, and healthcare costs.

**Summary:** In this series, we demonstrate that timely, short-term use of systemic corticosteroids among hospitalized patients with hypoxic respiratory failure due to SARS-CoV2 was well tolerated with good outcomes. The outcomes were reflected by reductions in inpatient mortality, CRP levels, requirement for mechanical ventilation and escalation of care.

## BACKGROUND

In December 2019, the novel coronavirus that causes Severe Acute Respiratory Syndrome was discovered in Wuhan, China. The previous epidemics caused by SARS-CoV (10% mortality) and MERS-CoV (37% mortality) caused more than 10,000 cases in two decades.^1,2^ As of June 18, 2020, there are more than 2,132,000 cases of SARS-CoV2 in the United States.^3^ The clinical spectrum of disease due to SARS-CoV2 continues to evolve and remains a challenge for physicians.

Most patients with mild illness tend to do well. However, patients with moderate or severe disease tend to be older, have obesity, have comorbid conditions such as diabetes mellitus, hypertension, and are more prone for hypoxic respiratory failure needing ICU level of care.^4,5^ The median time from onset of symptoms to hospital admission was five to seven days, and nine to ten days for ARDS and hypoxic respiratory failure.^6,7^ The pathophysiology involves an initial viral response phase where patients mostly have mild constitutional symptoms, followed by a pulmonary and then a hyperinflammatory phase where patients have shortness of breath, hypoxemia, abnormal chest imaging and elevated inflammatory markers.^8^

Current treatment for patients with SARS-CoV2 infection mainly involves supplemental oxygen, prone positioning along with supportive care. There are trials involving antiviral agents such as Remdesivir, hydroxychloroquine, and the use of convalescent plasma. However, a number of these trials mostly enroll patients in the early stage of the disease. Patients in the later stage (8 to 12 days) are often ineligible, leaving providers with few options to manage patients with worsening clinical and respiratory status.

A recent study from the UK showed that dexamethasone reduced deaths by one-third in ventilated patients and by one-fifth in COVID-19 patients that were on oxygen only.^9^ We postulate that early treatment with corticosteroids in patients with rising CRP and worsening hypoxemia may help prevent or attenuate the hyperinflammatory response associated with this condition. We report the clinical characteristics, early outcomes, and levels of C-reactive protein (CRP) in a limited set of patients with moderate or severe disease and received a short course of dexamethasone using our clinical management protocol.

## METHODS

This case series involves 21 patients with confirmed SARS-CoV2 who were seen by the Hospitalist team in Miriam Hospital, Providence, RI, USA between April 16^th^ and May 16^th^, 2020. A confirmed case was defined as a positive PCR assay for SARS-CoV2 in a nasopharyngeal sample tested by the laboratory at Miriam Hospital or Rhode Island Department of Health. The Institutional Review Board of the Hospital approved the study.

Patients were risk-stratified based on the severity of clinical illness, CRP levels, and oxygen requirements. Patients with mild disease had a minimal elevation of CRP (<50 mg/L), needed less than 2L/min oxygen, or only had GI symptoms. Moderate disease included patients with moderate elevation of CRP (50-200 mg/L) and 2-6L/min oxygen requirement. Patients with severe disease included patients with CRP greater than 200 mg/L and needed more than 6L oxygen requirement. Prone positioning was instituted in all patients with moderate to severe disease if they were able to tolerate it. Patients in the severe disease category were transferred to the Intermediate Care Unit for closer monitoring. They were also started on High Flow Nasal Cannula with or without Non-Invasive Ventilation. In addition, they were provided with an incentive spirometer, antitussives, and bronchodilators as needed.

Patients were offered short courses of dexamethasone at the discretion of their providers based on the following criteria and the KODA clinical management protocol (KODA was coined after one of the Clinicians at the Hospital):

1. Moderate disease with at least a 30% increase in CRP within 36 hours of admission, and increasing oxygen requirements.
2. All patients with severe disease with evidence of escalating oxygen requirements.
3. The presence of secondary bacterial infections as a probable cause of increasing CRP levels was excluded in all the selected patients.
4. Pulmonary embolism and cardiac dysfunction were excluded as probable causes of worsening hypoxia in all the selected patients.

Patients with the following set of criteria were excluded from the study:

1. All patients that showed significant clinical improvement and decrease in CRP levels and/or oxygen requirements on investigative modalities such as Remdesivir or Convalescent plasma.
2. Patient with associated COPD exacerbation who were primarily managed with systemic steroids.
3. Patients with Diabetic ketoacidosis, hyperglycemic hyperosmolar state, active concurrent bacterial infections.
4. Patients with a history of steroid-induced mania or psychosis.

Selected patients per the above criteria were administered dexamethasone 4mg three times daily for two days, followed by 4mg twice daily for two days and then 4mg once daily for two days.

The Infectious Disease service at Miriam Hospital followed these patients while they were admitted. Those who were eligible and were already enrolled in clinical trials for Remdesivir or convalescent plasma continued with their therapy. Hydroxychloroquine with or without azithromycin was given to patients who did not meet the criteria for the above trials and was showing signs of clinical deterioration and had no contraindications to the use of both medications. If the patient was observed to have a subsequent rise in CRP after initiating dexamethasone, the provider actively searched for secondary infection and promptly initiated broad-spectrum antibiotics. Corticosteroids were discontinued if there was no overt clinical benefit after 72 hours or clinical worsening after initiation.

### End Points

Our primary endpoints were escalation of care to the Intensive Care Unit (ICU) from General or Intermediate Care Level Unit, progression of respiratory failure needing mechanical ventilation and mortality. Secondary endpoints included change in CRP levels and Length of Stay (LOS).

### Data Collection

Data were obtained from the EPIC Electronic Medical Record system and recorded in a standardized case report form. Demographic data, Laboratory findings, Oxygen requirements in Liters Per Minute (LPM), Length of Stay, CRP levels, and Comorbid conditions were ascertained.

## RESULTS

We followed a total of 21 SARS-CoV2 positive patients. 61.9% of the patients were male, their mean age was 60 ± 15.77 years, mean BMI was 28.68 ± 9.46 kg/m^2^, and the most common comorbidities were Diabetes Mellitus and Hypertension (**Table 1**). The mean number of days after onset of symptoms to hospital admission was eight days. The mean maximum oxygen requirement was noted to be 15 LPM.

**Table 1:** Baseline Demographics of SARS-CoV2 Positive Patients.

| **Variables** | **Values** |
| --- | --- |
| Age (in years) |  |
| 30-49 | 28.57% (6) |
| 50-64 | 52.38% (11) |
| >65 | 19.04% (4) |
| Sex (Males) | 61.9% (13) |
| Race |  |
| White | 38.09% (8) |
| African American | 19.04% (4) |
| Hispanic | 28.57% (6) |
| Other | 14.28% (3) |
| Mean Onset of Symptoms (in days) | 8 ± 3.63 |
| Mean BMI (kg/m<sup>2</sup>) | 28.68 ± 9.46 |
| Comorbidities |  |
| Diabetes Mellitus | 38.09% (8) |
| Hypertension | 61.9% (13) |
| Coronary Artery Disease/Congestive Heart Failure | 14.29% (3) |
| Chronic Lung Disease | 14.29% (3) |

The mean CRP levels at the time of admission were 129.52 ± 72.05 mg/L. Peak levels were 185.09 ± 76.34 mg/L. At the time of discharge, mean levels were 40.73 ± 49.28 mg/L **(Table 2)**. There was 77.98% reduction in peak CRP levels after the initiation of dexamethasone. Dexamethasone was discontinued early in one patient due to hyperglycemia. None of the patients were transferred to the ICU or had progression of respiratory failure needing mechanical ventilation. All the patients required days of management in the Intermediate Level of Care Unit for their hypoxic respiratory failure using High Flow Nasal Cannula with or without Non-Invasive Ventilation.

**Table 2.** **Laboratory Values and Hospital Course of SARS-CoV2 Positive Patients**

|  |  |
| --- | --- |
| Mean CRP Levels on Admission (mg/L) | 129.52 ± 72.05 |
| Mean Peak CRP Levels (mg/L) | 185.09 ± 76.34 |
| Mean CRP Levels on Discharge (mg/L) | 40.73 ± 49.28 |
| Mean Length of Stay (in days) | 7.8 ± 3.82 |
| Mean Maximum Oxygen Requirement (Liters per minute) | 15 ± 9.16 |
| Antibiotic Use | 47.61% (10) |
| Concurrent Use of Investigational Therapies |  |
| Remdesivir | 42.86% (9) |
| Hydroxychloroquine +/- Azithromycin | 23.80% (5) |
| Convalescent Plasma | 66.67% (14) |
| Patients Discharged Home | 71.42% (15) |

Of the 21 patients that were admitted to the hospital, 71.42% were discharged home with a mean LOS of 7.8 days. Four patients were discharged to Skilled Nursing or Acute Rehabilitation Facilities. Two patients were transferred to inpatient hospice facilities on account of persistent hypoxemia, in line with their documented goals of care.

## DISCUSSION

In this limited case series, patients with rising CRP and worsening hypoxemia that received a short course of dexamethasone had a significant reduction in CRP levels and did not need to be transferred to the ICU or be mechanically ventilated. This may be a useful and low-cost tool that may help reduce critical care services utilization and free hospital beds, especially if there is a surge in inpatient volume.

SARS-CoV2 attaches to ACE2 receptors that are primarily located on type II pneumocytes. After infection, these cells release inflammatory signals that recruit macrophages, which in turn causes a “cytokine storm” that causes vasodilation, increased capillary permeability, and leucocyte migration. There is a release of Reactive Oxygen Species along with loss of surfactant, which causes the destruction of pneumocytes and the collapse of alveoli.^10,11^ This, in turn, leads to Severe Inflammatory Response Syndrome and further progression to Severe Acute Respiratory Syndrome in severe cases.

Early studies from China reported elevation of inflammatory markers including CRP, LDH, and IL-6 in patients with SARS-CoV2.^12^ CRP is a systemic marker of acute phase response in inflammation, tissue damage and infection.^13^ Elevated levels of CRP were associated with higher mortality, higher risk of clinical deterioration and progression of respiratory failure.^14^

Corticosteroids cause decreased vasodilation and permeability of capillaries, as well as decreased leukocyte migration to sites of inflammation. Glucocorticoids inhibit neutrophil apoptosis and demargination. They also inhibit NF-Kappa B, inflammatory transcription factors, and also promote anti-inflammatory genes like interleukin-10. They are known to inhibit the production of other mediators, including arachidonic acid metabolites such as cytokines, interleukins, adhesion molecules, and enzymes such as collagenase.^15,16^

Dexamethasone is a synthetic adrenal corticosteroid that has both anti-inflammatory as well as immunosuppressive properties. It is 20-30 times more potent than hydrocortisone and 4-5 times that of prednisone. Dexamethasone was chosen to reduce the need for diuretics to address fluid and sodium retention, which would have confounded interpretation of the results. In addition, dexamethasone has a longer half-life compared to other corticosteroids and will auto taper when discontinued.

Corticosteroids have always been controversial in the management of patients with Acute Respiratory Distress Syndrome (ARDS) and are not routinely used unless there is an alternate indication. Current guidance by the World Health Organization (WHO) recommends against the routine use of corticosteroids in patients with SARS-CoV2. Lee et al. showed that early steroid use in SARS-CoV1 patients resulted in prolonged detection of viral RNA and also did not benefit patients that received steroids later in their hospital course.^17^ On the contrary, Fang et al. showed that low dose corticosteroid administration did not effect viral RNA clearance.^18^ This can be explained by differences in the timing of initiation of therapy as well as patient heterogeneity. Numerous studies from China have reported routine use of corticosteroids in patients with SARS-CoV2, however, the risks and benefits are unclear.^6,19^ Two recent observational studies showed good outcomes in SARS-CoV2 positive patients with one study, even showing a mortality benefit.^20,21^ The trial by Villar et al. evaluated the use of dexamethasone in 277 patients with ARDS and demonstrated accelerated liberation from ventilation along with reduced mortality.^22^ Preliminary results from the RECOVERY trial showed that dexamethasone use prevented one death by treatment of around eight mechanically ventilated patients or approximately 25 patients requiring oxygen only. The trial studied the use of 6mg dexamethasone for ten days in contrast to our clinical management protocol that only used it for six days.^9^

A majority of our patients presented in the 7 to the 12-day time period with rapid clinical deterioration, consistent with progression to the hyperinflammation phase. The present study supports the use of corticosteroids during an optimal time window to help attenuate the severity of the inflammation and prevent a severe hyperinflammation phase. Two patients that received steroids much later in the course of their hospitalization did not significantly improve and were transitioned to inpatient hospice in line with their goals of care. Our study has many limitations. Being a case series, there is only a small number of patients that were followed. The study design has inherent biases along with patient selection. In addition, it is difficult to delineate the role played by other therapies administered concurrently along with corticosteroids. There is however, evidence that comparable patients who were enrolled in other therapies but did not receive the short course of dexamethasone ended up with a longer length of hospital stay.^21^

In conclusion, timely initiation of short course dexamethasone, a low cost, and relatively low-risk intervention may help prevent the progression of hypoxic respiratory failure in moderate to severely ill patients and help accelerate recovery. Further large-scale studies are urgently needed to study the role of early use of corticosteroids in SARS-CoV2 positive patients, especially those at high risk of clinical deterioration. Additional studies about the immune response associated with SARS-CoV2 infections, the use of velocity of CRP in disease monitoring, and the utility of dexamethasone with or without other therapeutic options are needed.

## Data Availability

The authors confirm that the data supporting the findings of this study are available within the article and/or its supplementary materials.

## ACKNOWLEDGMENT

We would like to thank the Infectious Disease Department at The Miriam Hospital.

